# Identifying robust biomarkers of infection through an omics-based meta-analysis

**DOI:** 10.1101/2020.07.28.20163329

**Authors:** Ashleigh C Myall, Simon Perkins, David Rushton, Jonathan David, Phillippa Spencer, Andrew R Jones, Philipp Antczak

## Abstract

A fundamental problem for disease treatment is that while antibiotics are a powerful counter to bacteria, they are ineffective against viruses. To ensure a given individual receives optimal treatment given their disease state and to reduce over-prescription of antibiotics leading to antimicrobial resistance, the host response can be measured to distinguish between the two states. To establish a predictive biomarker panel of disease state we conducted a meta-analysis of human blood infection studies using Machine Learning (ML). We focused on publicly available gene expression data from two widely used platforms, Affymetrix and Illumina microarrays, and integrated over 2000 samples for each platform to develop optimal gene panels. On average our models predicted 80% of bacterial and 85% viral samples correctly by class of infection type. For our best performing model, identified with an evolutionary algorithm, 93% of bacterial and 89% of viral samples were classified correctly. To enable comparison between the two differing microarray platforms, we reverse engineered the underlying molecular regulatory network and overlay the identified models. This revealed that although the exact gene-level overlap between models generated from the two technologies was relatively low, both models contained genes in the same areas of the network, indicating that the same functional changes in host biology were being detected, providing further confidence in the robustness of our models. Specifically, this convergence was to pathways including the Type I interferon Signalling Pathway, Chemotaxis, Apoptotic Processes, and Inflammatory / Innate Response. Amongst and related to these pathways we found three genes, *IFI27, LY6E*, and *CD177*, particularly prevalent throughout our analysis.

**Author summary:** Bacterial and viral disease require specific treatments, and whilst there are various treatment options for specific infection types, rapid diagnosis and identification of the optimal treatment remains challenging. Even in wealthier countries with developed healthcare systems, unnecessary prescription of antibiotics to patients with viral infections is causing phenomena such as multi-drug resistent bacteria. One way to distinguish a viral from bacterial infection is to measure an individual’s responses, for example by measuring the expression of particular genes in a blood sample, as different types of infections trigger different types of responses. In our study we analysed thousands of previously collected data sets from human blood, where individuals had either viral, bacterial or no infection (control). We used machine learning to identify “signatures” – small sets of genes that are indicative of the type of infection (if any) carried by an individual. Within data sets we used two different technology platforms had been used to collect data. We demonstrated that their gene-level signatures do not overlap perfectly when derived from the different platforms, the biological networks from which those genes were derived, however, had a high overlap – giving confidence that our models are robust against technology artefacts or bias. We have identified a small set of genes that serve as strong biomarkers of infection status in humans.

## Introduction

The varying differences within both classes of bacterial and viral infections cause the body to respond in a distinct way (1). Bacteria can be countered by pathways such as complement-mediated lysis, and the cell-mediated response for those that survive phagocytosis and live within the cell (intracellular bacteria). In this response, cells present bacterial peptides (antigens) on their surface, which are identifiable by Helper T cells that mediate bacterial destruction (2). There are a large variety of viruses and bacteria that affect the host’s immune system in various ways. Whilst some response pathways may overlap for bacterial and viral infections, there are however a number key differences (3, 4). In fact, these different response pathways cause varied transcription (expression) of key genes and are the medium for distinguishing disease state based on the host’s transcriptional response (5).

Differential expression of certain genes related to immunological responses can be indicative of both (i) disease state and (ii) individual pathogens (6). Such knowledge can be exploited in differentiating between viral, bacterial and control biological states. Previous studies demonstrated this by developing a small set of only seven genes that can accurately discriminate bacterial from viral infections across a range of clinical conditions, whilst simultaneously succeeding to determine with high accuracy which patients do not require antibiotics (7). Simultaneously, there have been numerous other studies looking at diagnosing infection based on the host’s transcriptional response (8-12).

Previous work failed to generalise as the data contains a far smaller set of pathogens that would be encountered in ‘real world’ scenarios, or studies focussed on single technology platforms, specific pathogens, or geographical regions (which contain populations with different HLA alleles, and different local pathogen groups). To address this lack of generalisation this work aims to utilise a larger scale analysis over a more representative sample set to improve biomarker generalisability. To gain statistical power and develop more robust panels, meta analyses of publicly available data have proven to be an effective technique (13). However, analysis integrating several cohorts together face inherent limitations from systematic variations otherwise known as “batch effects”. Without proper handling, these batch effects have been shown detrimental in population level gene expression analysis (14). Computational techniques exist to reduce batch to batch variation (15). ComBat (16), used in our study, is a well-known batch correction algorithm, and has been shown successful at removing batch effects between studies whilst retaining relatively high amounts of the biological variation.

Data-driven identification of robust biomarkers is a much-debated subject in the biological field. Several machine learning (ML) approaches have been proposed, with typically good performance on data sets used in a given study, but poorer performance when biomarkers are taken forward for validation. Important is the distinction between uni- and multi-variate approaches to biomarker discovery. While identifying a single predictive marker might be preferred in theory, multi-variate approaches have enabled the discovery of more complex relationships that can provide performance (accuracy; sensitivity) far exceeding univariate predictive models (17). One particular aspect in multi-variate predictive approaches is the optimisation of the representative model, which rarely can be achieved through brute force testing and relies on feature selection algorithms. In addition, models developed by ML approaches provide a more complete understanding of the underlying biological mechanisms, adding to our understanding of these systems. In this publication we focus on the use of the Random Forest (RF) (18) classifier, which has been demonstrated to perform well in real-world classification problems with high dimensionality and biased data (19). RFs are bagged decision tree models, which classify data points on a subset of features and have been praised for their ability to avoid overfitting (20). Unlike Support Vector Machines or Neural Networks (two frequently used models with high predictive capabilities) RFs forego much of the model selection step using an ensemble approach which builds many weak classifiers into a single strong self-averaging, interpolating model (21). Whilst RFs consist of many weaker models, they have been shown highly effective at capturing non-linear relationships between model predictors and outputs in a number of genomic studies (22, 23).

In recent years bioinformatician seeking predictive models have been faced with increasingly greater dimensionality to their data. With the needs of interpretable models many have responded and used feature selection procedures, which aims to remove redundant and irrelevant model features (24). The results of a smaller feature set not only offers improving model performance, faster computational implementation, and greater interpretation of the underlying generative process (25); but moreover lines up with the original pattern recognition theory, that RFs, like many other ML models were not designed to cope with large amounts of irrelevant features, often referred to as *the curse of dimensionality* (26). This high dimensionality is especially pronounced in the case of gene expression data with the total human gene set being ∼20,000.

Various feature selection procedure exist and have been demonstrated in biological problems (24). For this study we focused on Backwards Elimination (BW) (27) forming a well-established benchmark, and an evolutionary algorithm, a more explorative and parameterizable search approach, to obtain reduced model feature sets (17). BW essentially searches for the optimal feature set by progressively eliminating the least important features from a given dataset and testing whether the new model is significantly more accurate than the previous. Whereas evolutionary algorithms are based on evolving population(s) of models, which are repetitively intermixed, and subject to random point mutations. This evolutionary process is assumed to produce converging model populations in terms of performance and their associated feature sets (28).

The application of different computational pipelines often leads to different outcomes in disease prediction (29). We believe, it is thus important not only to present performance statistics for one given model generated by an ML pipeline, but to explore the underlying biological response of a set of plausible models. By doing so, it is possible to develop a more robust biomarker panel (mitigating overfitting which would generally produce models hard to interpret biologically), and to understand why a given model, or set of similar models, are valid.

In this work, we have performed a meta-analysis over publicly available transcriptomics data (human blood samples where individuals had bacterial, viral or no infection), from two microarray technologies (Affymetrix and Illumina). We applied feature selection and machine learning for biomarker discovery and predictive model generation, and lastly we explored the biological context of the resulting models by reverse engineering the underlying networks. Representing omics data as a network, has several key benefits. One can often better represent many complex systems as connected components, and the genome is no exception (30). Clustering is one popular method to explore these complex networks and many algorithms exist to reveal insight into these complex structure (31). Visualising a clustered network allows us to explore aspects of this generative process, and how feature selection unfolds over it. However, network construction can often be sensitive to the computational approach and parameterization applied (32, 33). In our approach, we validated our findings and mitigate any potential bias in network generation and clustering by illustrating that the biological driven feature selection is consistent across two separate networks, containing different studies, and derived from different technological platforms.

## Materials and Methods

To identify and validate a panel of biomarkers able to differentiate bacterial and viral infections, we performed a meta-analysis of GEO gene expression data, all from open source microarray human blood infection studies. Our analysis was divided into three major method steps: i) pre-processing, ii) feature selection, and iii) inferring a gene interaction network, to discover and our validate gene lists (Fig 1). Following the major steps, we performed and report the results of a final out of sample test on data not previously used in the training phase for greater validation.

**Fig 1.**
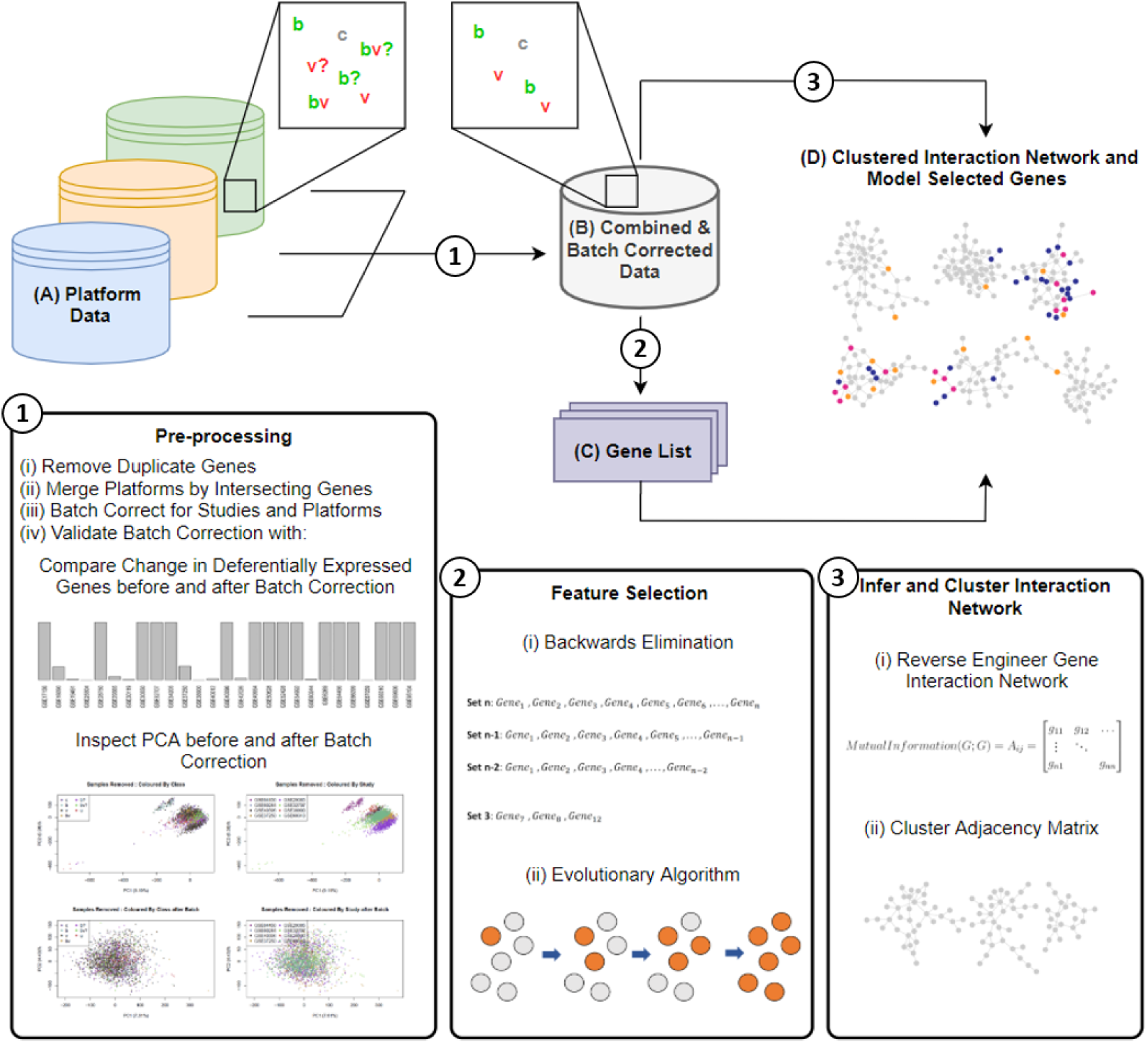
Conceptual overview. Individual data (A), containing bacterial (b), viral (v), control (c), and samples with lower levels of study confidence (?s) is merged by common genes and pre-processed by Step 1. Step 1 outputs a combined and batch corrected dataset (B), where only b/v/c samples are present. Two instances of (B) are formed, one where samples of lower levels of support are integrated into b/v classes, and the other completely omitting uncertain samples. Feature selection is performed on data B in Step 2 using (i) Backwards Elimination, and (ii) an Evolutionary algorithm. Step 2’s output is a number of Gene Lists (C) obtained in the feature selection. Data B is also used to infer and cluster a gene interaction network, by (i) reverse engineering the gene interaction network, and (ii) clustering the adjacency matrix. (D) is then formed as the clustered interaction network overlaid with genes found in the best performing mode of each dataset and search procedure.

### Pre-processing

#### Data

Datasets from four technological platforms (two from Affymetrix platforms and two from Illumina platforms), consisting of 3868 samples, from 21 different studies, were included in the analysis (Table 1). These datasets were selected from a wider pool identified in an initial scan of online databases, based on a variety of factors including: microarray platform manufacturer (most prevalent platforms – Affymetrix and Illumina) and study set size (larger studies with more predictive power), class pathogen strain distribution (aiming for an equal distribution across the data); and ability to merge with other datasets in our analysis.

**Table 1.**
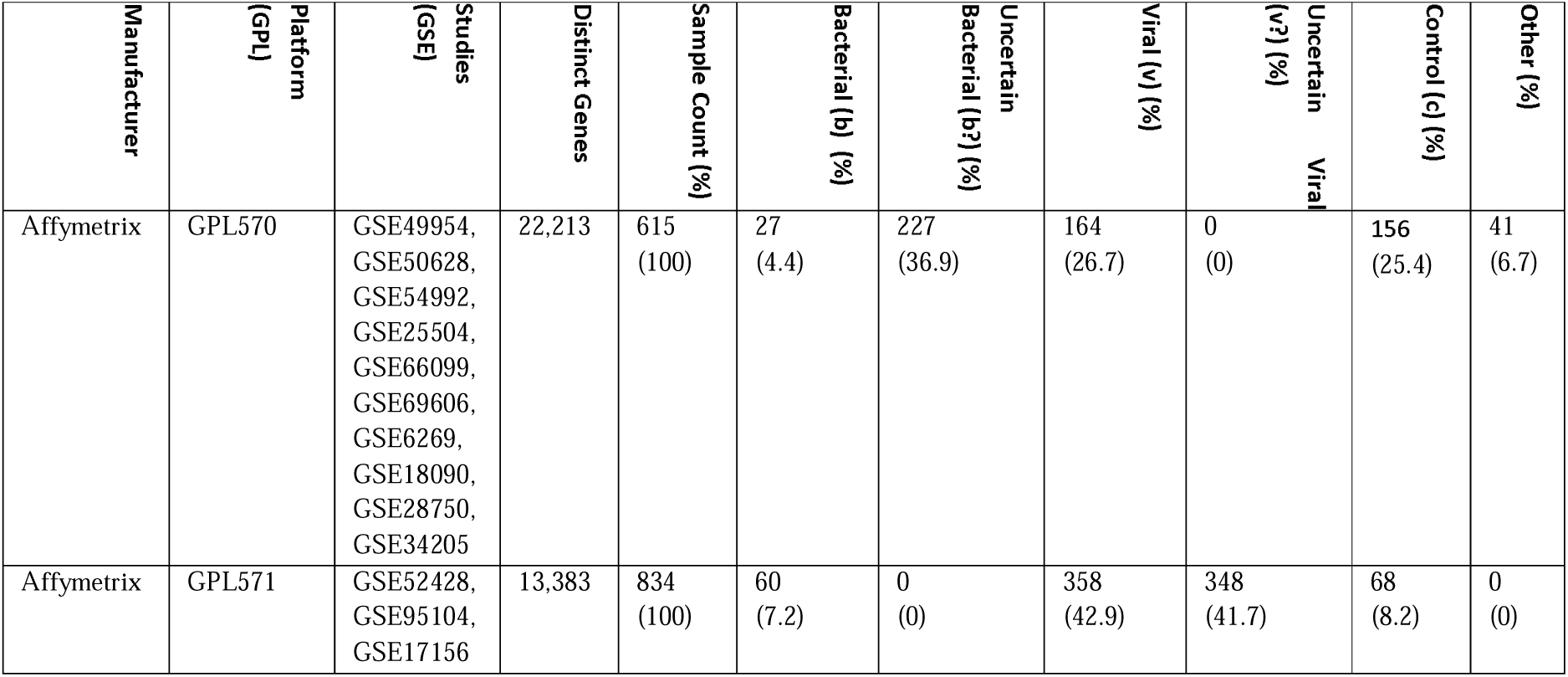
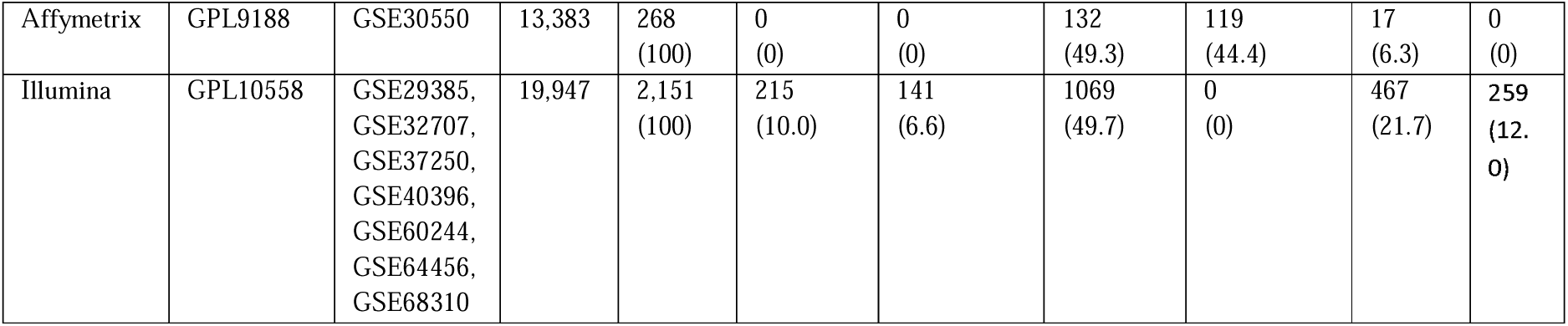
Summary of platform level Affymetrix and Illumina datasets prior to pre-processing.

| Manufacturer | Platform (GPL) | Studies (GSE) | Distinct Genes | Sample Count (%) | Bacterial (b) (%) | Uncertain Bacterial (b?) (%) | Viral (v) (%) | Uncertain Viral (v?) (%) | Control (c) (%) | Other (%) |
| --- | --- | --- | --- | --- | --- | --- | --- | --- | --- | --- |
| Affymetrix | GPL570 | GSE49954, GSE50628, GSE54992, GSE25504, GSE66099, GSE69606, GSE6269, GSE18090, GSE28750, GSE34205 | 22,213 | 615 (100) | 27 (4.4) | 227 (36.9) | 164 (26.7) | 0 (0) | 156 (25.4) | 41 (6.7) |
| Affymetrix | GPL571 | GSE52428, GSE95104, GSE17156 | 13,383 | 834 (100) | 60 (7.2) | 0 (0) | 358 (42.9) | 348 (41.7) | 68 (8.2) | 0 (0) |
| Affymetrix | GPL9188 | GSE30550 | 13,383 | 268<br>(100) | 0<br>(0) | 0<br>(0) | 132<br>(49.3) | 119<br>(44.4) | 17<br>(6.3) | 0<br>(0) |
| Illumina | GPL10558 | GSE29385,<br>GSE32707,<br>GSE37250,<br>GSE40396,<br>GSE60244,<br>GSE64456,<br>GSE68310 | 19,947 | 2,151<br>(100) | 215<br>(10.0) | 141<br>(6.6) | 1069<br>(49.7) | 0<br>(0) | 467<br>(21.7) | 259<br>(12.0) |

#### Deduplicating genes by probes and merging datasets

Dataset columns (originally microarray ProbeIDs) were first deduplicated and substituted by their gene mappings. Where duplicate ProbeIDs existed for the same gene we selected a representative ProbeID with the highest average intensity across samples (34). Samples from datasets of the same manufacturer were then merged by common genes, first at the level of studies within the same platform, then by platforms in the same manufacturer.

#### Batch correction and evaluation

Batch corrections targeted two non-biological sources of systematic variation: (i) inter-platform study batch effects (differences between platforms), and (ii) intra-platform batch effects (differences between studies within a batch). Batch correction was implemented with ‘ComBat’ (16) in a two-step sequential batch correction pipeline (S1 Appendix.docx). We repeated this process for both Affymetrix and Illumina datasets separately to form batch corrected Affymetrix data, and batch corrected Illumina data.

Batch correction was verified to retain biological variation and remove technical variation using two validation steps (Fig 1 Step 1). Firstly, we tested whether pre and post batch correction significant features overlapped significantly. Secondly, we performed Principle Component Analysis (PCA) (35) visualising the data in two dimensions and comparing the PCA plots of before and after batch correction. For a successful batch correction, pre-batch correction sample clustering in the PCA would be visually removed in the PCA plot of post batch corrected data.

#### Dealing with study sample ambiguity: forming a confirmed and integrated dataset instance

To include more data, including some class ambiguity in the original studies, we formed a modelling dataset which integrated bacterial and viral samples with lower levels of confidence (b?, and v?)(Table 1). This integrated dataset contained only classes labeled b/v/c (Fig 1). For Affymetrix this formed (Affy_I) and similarly, for Illumina this formed Illumina_I. Two additional datasets of confirmed sample classes only, were also generated and included in the study but presented only in the Appendix.

#### Feature Selection

To search for optimal panels of genes we implemented two search feature selection procedures: (i) the well-known Backward Elimination process (27), and (ii) a genetically inspired search algorithm (GALGO) (17). Both search procedures operated using the RF Classifier, implemented in the R Ranger package (36) a fast and parallelisable implementation of RFs for high dimensional data.

#### Dataset Preparation

For dealing with un-even class distributions present in our data (Table 1) we employed two strategies. Firstly, we used a study aware data split which insured relatively equal class proportions across both training, test and evaluation data splits. Secondly, we ensured that classification accuracy bias due to larger class proportion of disease states was minimized by weighing smaller classes correspondingly higher (18). This ensures that our model will not be biased to classifying samples with a larger proportion in the dataset.

#### Backward Elimination

We operated on a 60/20/20 training/test/evaluation data split for each dataset processed in BW (37). On each training set we ran 240 BW search procedures, using Out-of-bag (OOB) error as the minimisation criterion and implementation using the VarSelRF R package (38). Each run generated a single optimal model which minimised OOB. For each dataset a single representative model was selected from the 240 runs which maximised accuracy on test data.

#### Genetic-algorithm

The Genetic-Algorithm (GA) optimized approach is an efficient method for creating suitable multivariate models. We used the R library GALGO (17) to identify a small feature model by continuously crossing a number of small feature models (chromosomes of features) with each other, hypothetically identifying better models with successive generations. We used an initialised fitness goal of 0.95, model size (chromosome size) of 15 genes, and k-fold cross-validation to counter overtraining. In the RF, larger classes, namely viral, were also penalized, as to ensure equal predictions across classes. After 250 models, we generated a representative model through a frequency based forward selection strategy which ensures only genes that contributed to predictions are included in the final model (S2 Appendix).

#### Inferring underlying interaction network

We reverse engineered gene regulatory networks using ARACNe (39) which builds an adjacency matrix of genes with their mutual information from expression data (Fig 1). These networks allow identification of functional relationships between genes and their corresponding products (40, 41). In addition, they can provide insight into the functionally relevant groups of genes for distinguishing disease state, by examining locations of RF selected genes.

To select significant interactions within our dataset we used a p-value threshold < 0.05 in the ARACNe procedure. The approach can then estimate a mutual information threshold that is relevant for the provided dataset and a specified p-value. With our data this resulted in a threshold of MI > 0.0176 to be retained. From the gene pairs of mutual information, we formed an edge table which was the basis for our interaction network. Nodes are genes and edge weights are the mutual information between two genes, where greater mutual information would suggest a stronger relationship. We then loaded our networks in Cytoscape (42) which visualises molecular interaction networks and has support for a number of clustering algorithms.

To identify highly interconnected sub-networks within our reconstructed regulatory network we utilised the Cytoscape clustering plugin GLay (32). GLay uses an implementation of the Girvan-Newman Edge-betweeness algorithm (43) which we used to split our networks it into clusters of connected genes. This resulted in a number of smaller sub-networks and allowed us to inspect their functional roles within the larger network. We then mapped higher level ontologies, such as pathways and gene ontology from gene symbols and used the DAVID (44) tool to provide enrichment analysis. The enrichment analysis looked at several different ontologies, providing an indication of overrepresentation, which we used to infer the likely biological function of a given cluster. Each cluster analysis generated an enrichment table detailing enriched ontology terms along with enrichment ratios and (adjusted) p-values. From the enrichment table we then produced a dotplot which depicted enrichment ratio, p-value and gene count, along with a colour scheme denoting different ontologies, for visual interpretation.

For clusters of genes with enriched and significant terms related to the immune response, we labelled them manually as Functionally Relevant (FR) clusters. These FR clusters allowed us to make inferences about which biological functions hold predictive power, by overlaying model selected genes onto our labelled gene regulatory network.

#### Out of sample testing

Out of sample testing usually refers to testing a model on data not previously seen in model training and selection (37). Whilst a validation set was held back for both Affymetrix and Illumina data, the validation data still contained samples from the same manufacturer and group of studies used in training. Hence, within the original ‘discovery dataset’, gene lists could still be overfit to some non-biological effect persisting in either the manufacturer technology or set of studies present, which was not removed by batch correction.

To properly test generalisability and investigate any discovery data bias, we evaluated the best performing models discovered on both Affymetrix and Illumina data by retraining and testing them on non-discovery data (Affymetrix Gene Lists to Illumina Data, and Illumina Gene lists to Affymetrix Data). These non-discovery datasets contained samples from different studies and technology and therefore represented the ideal validation datasets. With similar error between discovery and non-discovery data one can be confident that models have not overfitted to a given dataset and are suggested to be generalisable.

## Results

### Pre-processing

Gene de-duplication and data merging was successful for both Affymetrix and Illumina. In the final Illumina datasets 19,947 distinct genes were found intersecting all studies, whereas for Affymetrix Data we found 13,383 (Table 2). This lower Affymetrix count was due to platforms GPL571 and GPL9188 having only 13,383 distinct genes (**Table 1**). This gene loss from intersection resulted in the omittance of 8,830 gene columns, which were present for the 615 samples in GPL570.

**Table 2.**
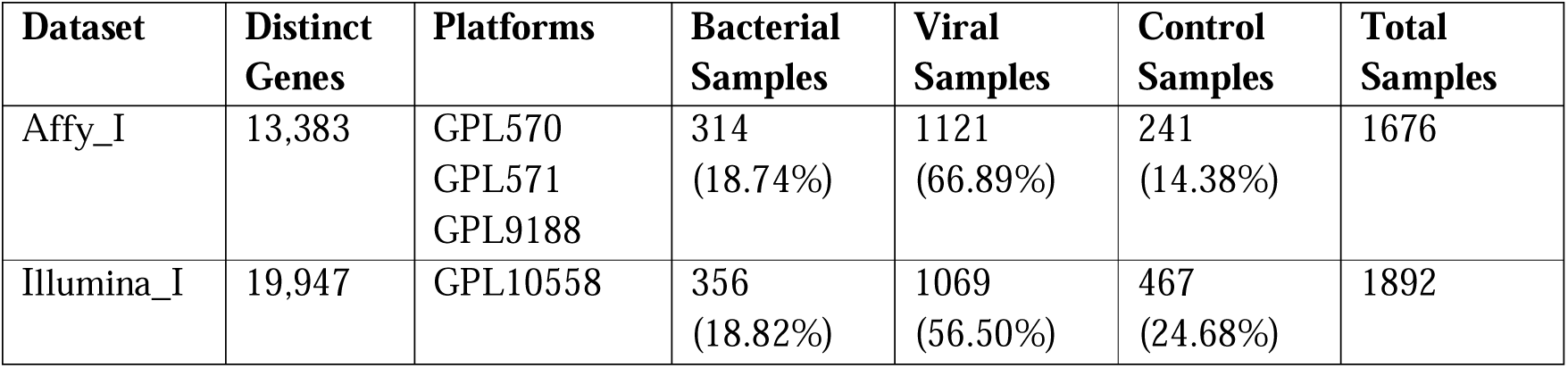
Merged and batch corrected modelling dataset description. Merged and batch corrected Affymetrix and Illumina (ambiguous classes integrated) dataset breakdown by distinct genes, platforms, class make up, and sample count.

| Dataset | Distinct Genes | Platforms | Bacterial Samples | Viral Samples | Control Samples | Total Samples |
| --- | --- | --- | --- | --- | --- | --- |
| Affy_I | 13,383 | GPL570<br>GPL571<br>GPL9188 | 314<br>(18.74%) | 1121<br>(66.89%) | 241<br>(14.38%) | 1676 |
| Illumina_I | 19,947 | GPL10558 | 356<br>(18.82%) | 1069<br>(56.50%) | 467<br>(24.68%) | 1892 |

Affymetrix platforms were successfully merged and combined via our batch correction pipeline, indicated by non-significant changes in differentially expressed (DE) genes and removal of clustering in our PCA analysis between both study and platform batch corrections (S1 Appendix). Illumina based datasets were represented by a single platform, GPL10558. Batch correction did not result in significant changes to DE genes and removed the previously observed clustering by study (S Fig 2).

**Fig 2.**
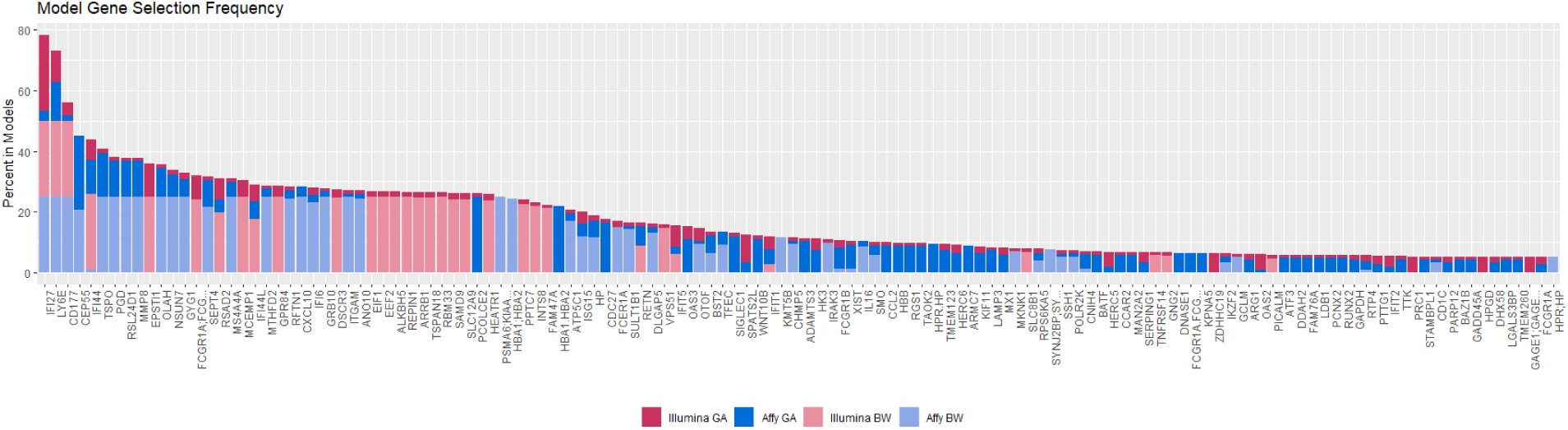
Gene frequency in Affymetrix and Illumina models. Each Model frequency is scaled between 1 and 25. Model overlapping gene frequencies are then stacked and coloured by model-dataset combination. Affymetrix Models by shades of blue and Illumina models by shades of red.

This resulting two datasets Affy_I and Illumina_I contained 1676 and 1892 samples respectively (**Error! Reference source not found**.). It is evident there is an uneven class distribution present in both datasets. Both Affy_I and Illumina_I are made up of more than 50% viral samples (66.89% and 56.50%, Table 2). The most underrepresented class is bacterial samples, with both datasets comprising fewer than 20% samples labelled as bacterial (Table 2).

### Biomarker lists

Running GA and BW on both Affymetrix and Illumina generated an ensemble of models for each method-datasets pair. For BW this was an ensemble of optimal models, one per run of the algorithm. For GA this was the evolved chromosomes obtained by repeats of the search procedure. From this ensemble of models, we computed relative gene selection frequencies (top 16 genes displayed Table 3).

**Table 3.**
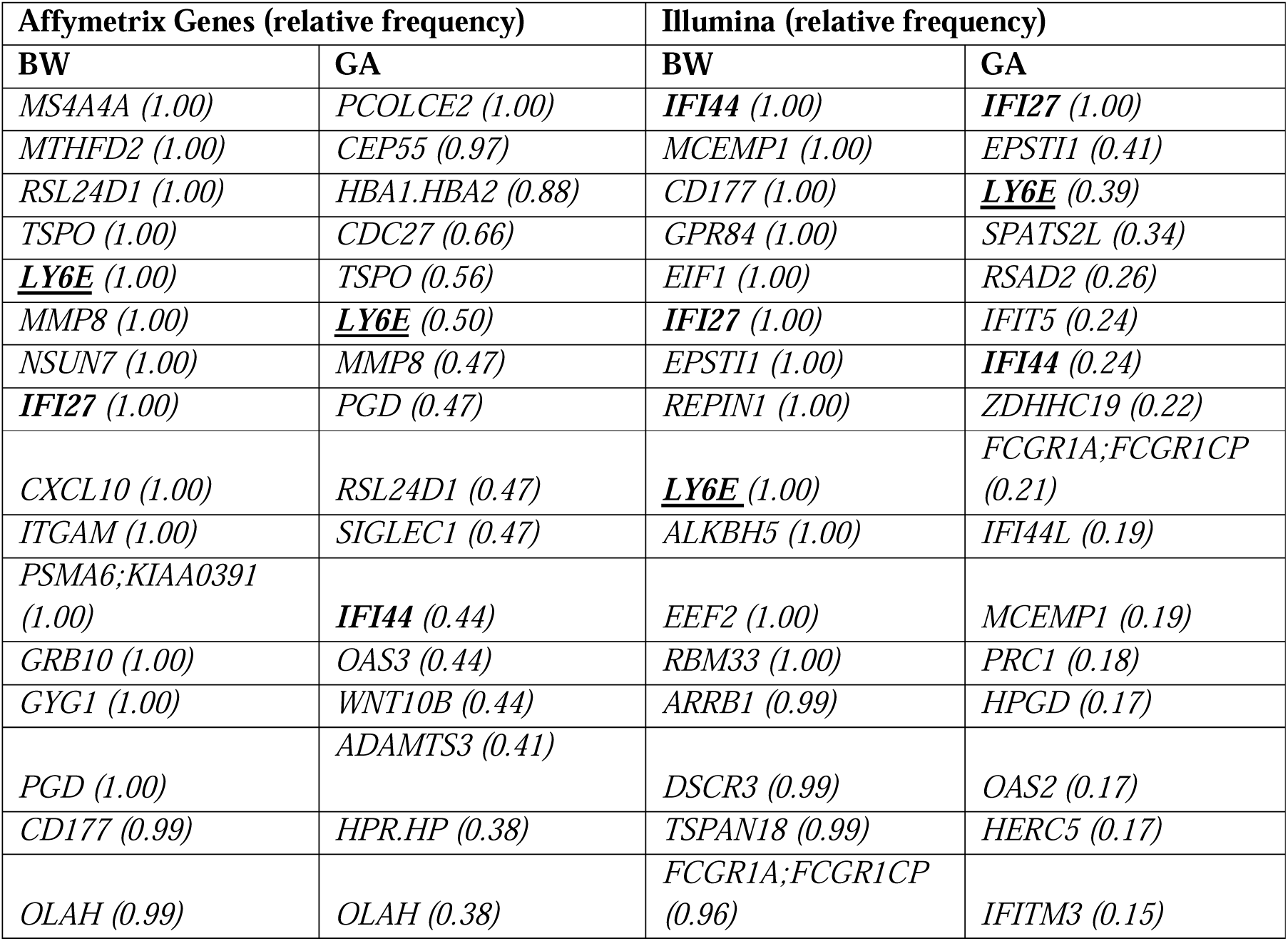
Top 16 Gene selection for Affymetrix and Illumina models and their relative selection frequencies. Frequency provided in brackets is based on the model selection frequency in each optimisation run (the number of times a gene was selected across the number of optimised models). Bold genes are included amongst 3 of models top 16 selection, and underlined genes are included in all four.

BW search procedures in both technologies converged to a small set of genes, indicated by high relative selection rate calculated by the number of times a gene was selected across the multiple runs performed in each optimisation procedure. For Affymetrix 14 were included at a rate of 1.0, whereas for Illumina BW results contain 12 genes at a rate of 1.0 (Table 3). GA’s on the other hand contained a much wider gene selection in the evolved chromosome, in both search procedures only a single gene was included at a relative rate of 1.0. which reflects the more varied selection in GA search procedures.

Overall search results (aggregated between runs by frequency) from BW and GA in both Affymetrix and Illumina all contained *LY6E* (Lymphocyte antigen 6E, UniProt: Q16553) amongst their 9 most frequently selected genes (Table 3). Amongst the next widely selected genes were *IFI27* (Interferon alpha-inducible protein 27, mitochondrial, UniProt: P40305) and *IFI44* (Interferon-induced protein 44, UniProt: Q8TCB0), both in the top 16 by gene selection frequency for three of the four search procedures (Table 3). These 3 genes (*LY6E, IFI27*, and *IFI44*) are all type-I interferon-inducible genes (ISGs), demonstrated to have altered expressions in disease states, and known to be highly effective at countering infection (45-48). Furthermore, an additional number of other ISGs were also found amongst the frequently selected model genes (*MS4A4A, IFI44L, OAS2, and IFIT5*).

Additionally, several other most frequently selected genes have been linked to certain disease states in the literature. Particularly increased levels of *MMP8* have been observed in HIV-infected patients, which cross-references well as a high proportion of samples in our modelling data coming from HIV viral studies (49). *SIGLEC1* is a Type I transmembrane protein expressed by a subpopulation of macrophages and was one of fifteen genes found upregulated during *in vivo* respiratory syncytial virus infections (50), whilst also said to initiate the formation of the virus-containing compartment (51).

To further investigate gene convergence, we compared the relative model gene inclusion rates for all search procedures together. We scaled each model gene frequency (between 1 and 25), then plotted them together as a stacked bar plot. Fig 2 shows the resulting stacked frequency, where genes are visualised for greater than 5% aggregated inclusion across all search procedures. Similarly, to our top 16 gene comparison, *LY6E* is indicated as important, being represented in all search procedures. However, interestingly *IFI27* is also included amongst all search procedures. Furthermore *CD177*, a neutrophil-specific receptor and known to be at increased expression for patients in septic shock (52, 53), was selected relatively frequently and present in all search procedures.

One interesting aspect to look at is the intersection of this between Genes frequently selected between Affymetrix and Illumina generated models. We identified 88 genes intersecting between Affymetrix and Illumina (S1 Table) and performed functional enrichment analysis of them using DAVID. We found both highly enriched and significant terms relating to the immune response. Included in the list of significant pathways was, in order of significance

For each search procedure we obtained a final representative model (Affy_BW, Affy_GA, Illumina_BW, and Illumina_GA) and evaluated its performance on a held-out data split. Model performance was recorded as the size of the gene list and its class-based performance in terms of: Balanced Accuracy, Sensitivity, Specificity, and Mcnemar’s Test p-value which tests for consistency in responses and can reveal bias to classifying a certain class (all metrics derived from the evaluation data split) (54).

Average model size was similar between both Affymetrix and Illumina models (30-37 genes) (Table 4). On average models classified 0.89 of Bacterial, 0.72 of Control and 0.86 of Viral classes correctly across all datasets. In particular, the Affymetrix models, BW and GA, performed particularly well in terms of balanced accuracy on bacterial samples (0.94 and 0.93 respectively). In terms of sensitivity all models performed well for bacterial and viral classes (on average 0.85, and 0.93 respectively), however control sample performance was worse when compared to the viral and bacterial classes (0.57). Evaluating model specificity, bacterial performance was particularly high over all models (averaging 0.95) which would suggest we can determine what a bacterial sample is particularly well regardless of the model used (Table 4).

**Table 4.** Overall optimal model performance. Model performance break down by Affymetrix and Illumina data sets on the held out test dataset in terms of final model gene size, Balanced Accuracy, Sensitivity, Specificity, and Mcnemar’s Test p-value.

|  | Model-Dataset<br>Combination | Gene-<br>set<br>Size | Balanced<br>Accuracy<br>(B/C/V) | Sensitivity<br>(B/C/V) | Specificity<br>(B/C/V) | McNemar’s<br>Test p-value |
| --- | --- | --- | --- | --- | --- | --- |
| Affymetrix<br>Identified<br>Models | BW | 33 | 0.94 / 0.78 /<br>0.86 | 0.90 / 0.57 /<br>0.97 | 0.93 / 0.96 /<br>0.76 | 3.57e-3 |
|  | GA | 36 | 0.93 / 0.82 /<br>0.89 | 0.88 / 0.66 /<br>0.97 | 0.99 / 0.97 /<br>0.81 | 4.90e-10 |
| Illumina<br>Identified<br>Models | BW | 30 | 0.86 / 0.70 /<br>0.78 | 0.80 / 0.47 /<br>0.87 | 0.93 / 0.92 /<br>0.87 | 2.36e-3 |
|  | GA | 37 | 0.82 / 0.58/<br>0.89 | 0.83 / 0.58 /<br>0.89 | 0.93 / 0.94 /<br>0.77 | 4.33e-15 |
| Average | Average | 34 | 0.89 / 0.72 / | 0.85 / 0.57 / | 0.95 / 0.95 / | 5.93e-3 |
|  |  |  | 0.86 | 0.93 | 0.80 |  |

### Inferred interaction networks

We inferred the underlying gene regulatory networks for both Affymetrix and Illumina datasets, but present here the analysis on the larger Illumina network (Affymetrix analysis in S3 Appendix). GLay clustering of the gene interaction network initially revealed 14 clusters containing more than 10 genes (Fig 4). To enable a more granular analysis of specific network sections (those indicated to be functionally relevant in the immune response (FR) as indicated by enrichment analysis, or containing genes selected by our models) we further partitioned several of the initial clusters, forming a network hierarchy (limited to a depth of 3). This resulted in 110 distinct groups of genes which we analysed (Table 5).

**Table 5.** Illumina interpreted inferred interaction network properties. Clusters have been labelled either functionally related to the immune response (FR). For a cluster to be labelled as FR, functional enrichment analysis of their gene list will have revealed terms both enriched and significant implicated in the host response to disease.

| Manufac<br>turer | Nodes<br>(Genes) | Sub-clusters<br>of more than<br>4 Genes | FR<br>Clusters<br>(% of all) | FR Clusters<br>selected by > 1<br>Model (% of all) | FR Clusters<br>selected by all four<br>Models (% of all) |
| --- | --- | --- | --- | --- | --- |
| Illumina | 19839 | 110 (1.00) | 24 (21) | 10 (9) | 4 (4) |

**Fig 3.**
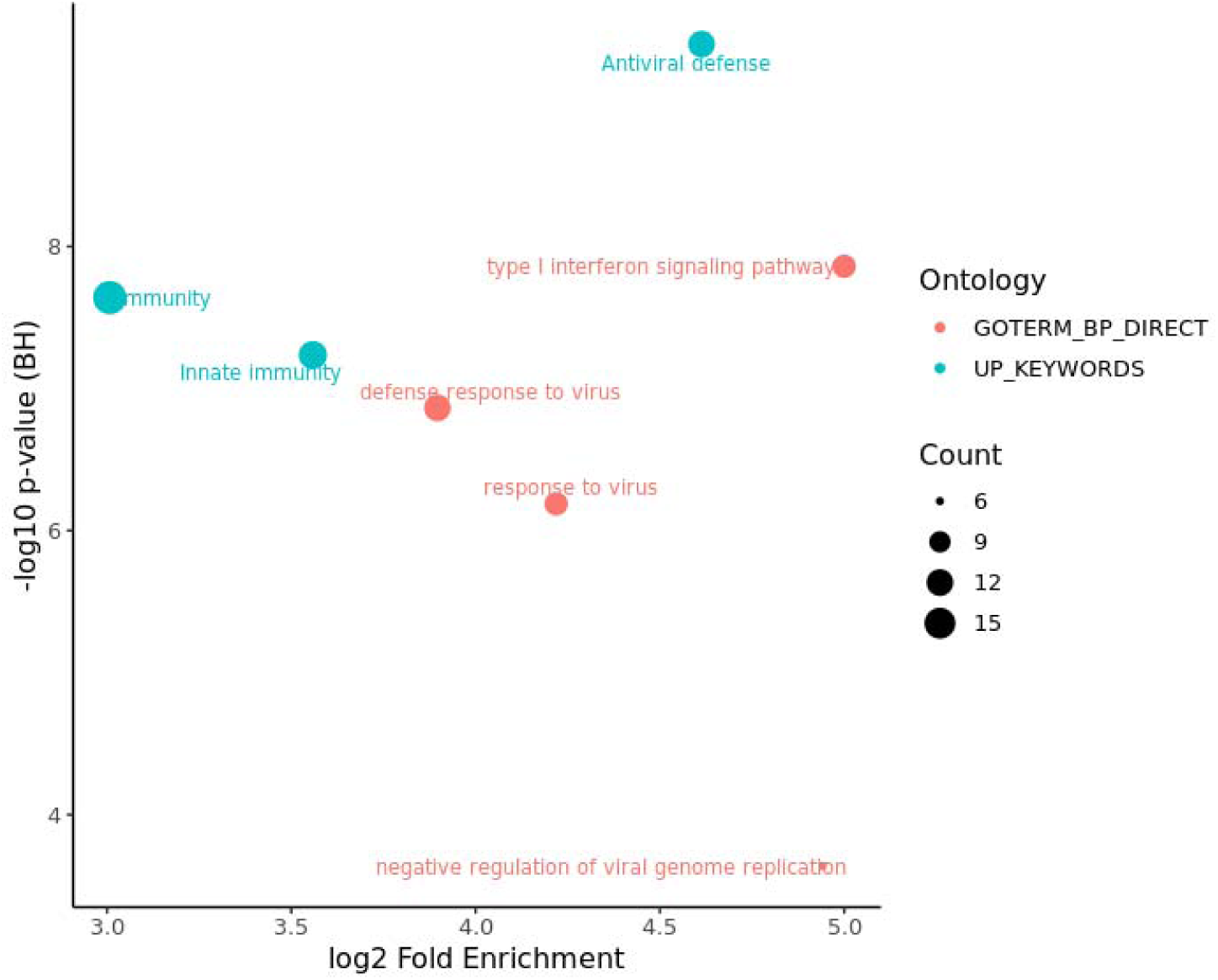
Functional enrichment analysis of the identified 88 genes intersecting between Affymetrix and Illumina search procedures. ‘Antiviral defense’ is the most significant term, whilst ‘type I ‘Antiviral defense’ comprising of 12 genes, the ‘type I interferon signalling pathway’ which included 10 genes, and ‘Immunity’ encompassing 17 of the 88 genes intersecting between Affymetrix and Illumina search procedures (**Fig 3**).

**Fig 4.**
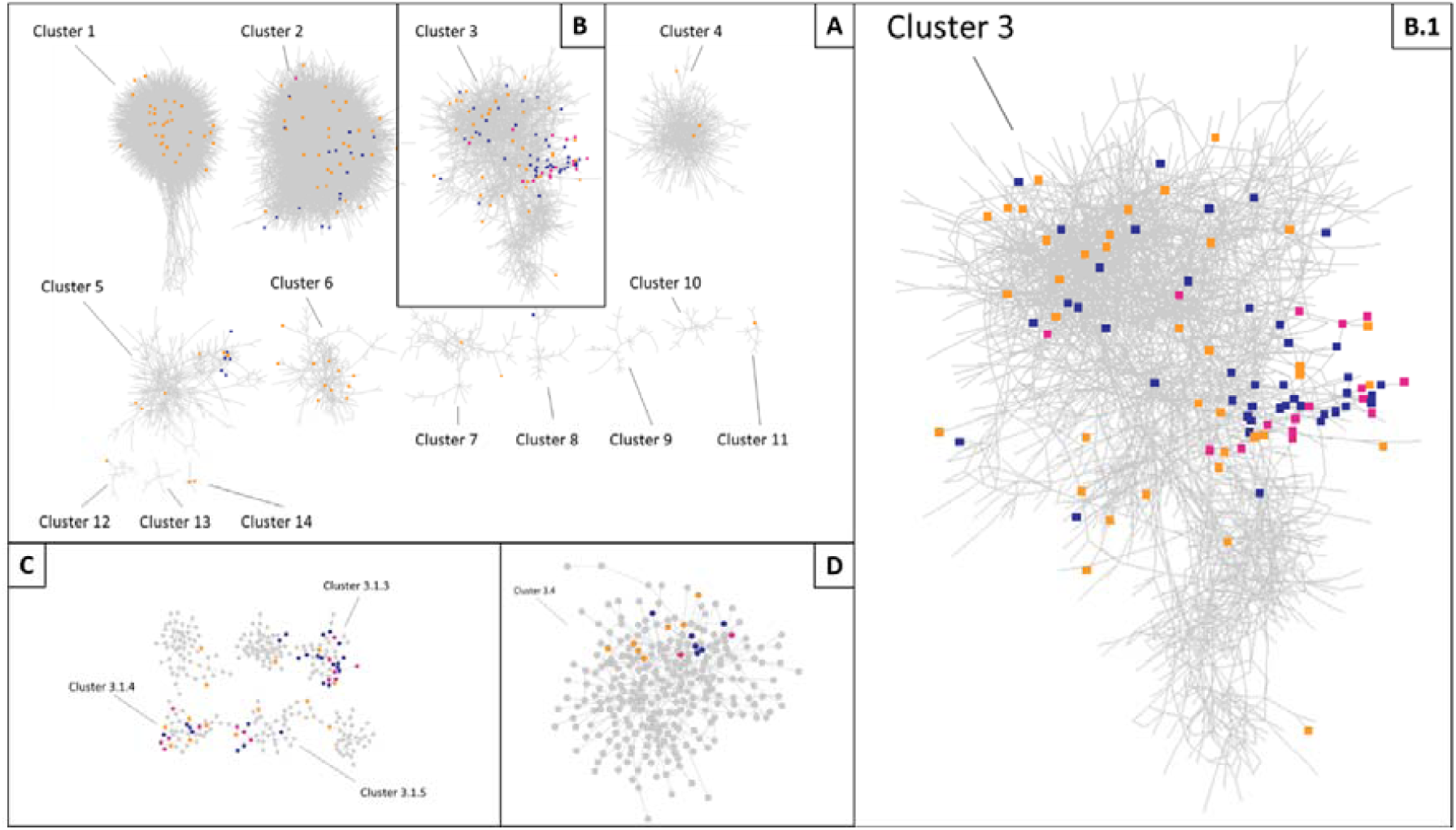
Clustered Illumina interaction network. Illumina models’ selected genes are blue, Affymetrix selected genes are orange, and those intersecting both technologies are pink. (A) Illumina Interaction network after initial clustering (visualising clusters > 10 Genes). (B) Cluster 3, containing the most selected genes which intersected between Affymetrix and Illumina models. (B.1) Cluster 3 Enlarged. (C) Highly selected sub clusters of Cluster 3. (D) Cluster 3.4, a sub cluster of Cluster 3 containing two genes which were selected by both Affymetrix and Illumina models.

In Illumina 24 of the 110 clusters had enriched and significant terms related to functions of the immune system in our DAVID analysis (Table 5). Of these 24 FR clusters, 10 had at been selected by at least one Illumina model. These 10 clusters contained 55 genes in the union of Illumina models (68% of all 81 Illumina selected genes in the network). Additionally, a small number of clusters (four) were selected by every model.

### Affymetrix – Illumina cluster comparison

We found a similar number of clusters converged to between both Affymetrix and Illumina gene lists in their respective networks (S3 Appendix). It is clear the RF models are selecting genes from multiple of these uncorrelated clusters, to build a stronger, less correlated model feature sets able to define disease state. For greater biological understanding we compared the most selected clusters from both the Affymetrix and Illumina Interaction Network. In Illumina this was Cluster 3.1.3 (S3 Appendix). Whilst the size between both clusters was not comparable (Affymetrix – Cluster 5 being 435 Genes and Illumina Cluster 3.1.3 being only 47) we found an intersection of 16 Genes (*DDX60, IFI35, IFI44, IFI44L, IFIH1, IFIT1, IFIT2, IRF7, ISG15, MX1, OAS2, SCO2, TIMM10, TRAFD1, TRIM22 and ZBP1*) which was statistically significant (p-value < 3.18e-12), 10 of which known to be ISGs (*IFI35, IFI44, IFI44L, IFIH1, IFIT1, IFIT2, IRF7, ISG15, MX1, OAS2)* (47). Performing DAVID enrichment analysis on both clusters, we find in Illumina Cluster 3.1.3 one highly enriched term ‘type I interferon signalling pathway’ albeit with a non-significant p-value (S3 Appendix). We do not see the same term in the Affymetrix cluster; however, it does contain numerous ISGs, which we saw commonly amongst gene lists. This convergence between independent feature selection across separate manufacturers and different studies reinforces the high predictive power of ISGs for discriminating disease state across infection studies.

### Independent cluster convergence between Affymetrix and Illumina models

To examine whether convergence between Affymetrix and Illumina was also to the same clusters containing the same genes we looked at where in the Illumina interaction network Affymetrix gene lists selected from (Fig 4, full break down in S3 Appendix). Although selected genes varied between Affymetrix and Illumina, we indeed found that both converged around the same clusters of genes. Moreover, we found that 19 clusters (including lower level sub clusters) were selected by both Affymetrix and Illumina models in the Illumina interaction network. Interestingly amongst this set, the four sub clusters intersecting across all Illumina gene lists (all from within the larger Illumina-Cluster 3: Fig 4) were also selected by Affymetrix gene lists: Illumina-Cluster 3.1.3, Illumina-Cluster 3.1.4, Illumina-Cluster 3.1.5, and Illumina-Cluster 3.4. All of these clusters contained genes revealed by selection frequency analysis in previous section 4.2.

We investigated all four clusters selected by all Illumina models (Clusters 3.1.3, 3.1.4, 3.1.5 and 3.4) and found they could be separated functionally to different aspects of an immune response. As mentioned, enrichment analysis on Illumina Cluster 3.1.3 revealed the ISGs to be present. However, enrichment analysis also revealed a number of both highly enriched and significant terms related to viral infections (‘response to Viruses’, ‘defense response to virus’), and most prominently ‘Antiviral Defense’ which is no surprise given the high number of interferon related genes in the cluster (S3 Appendix). Comparing the 47 genes in Clusters to our model frequency analysis revealed 18 overlapping genes (*DHX58, EPSTI1,HERC5, IFI44, IFI44L, IFI6, IFIT1, IFIT2, IFIT5, ISG15, MX1, OAS2, OAS3, RSAD2, RTP4, SAMD9, SPATS2L, and TMEM123*).

For cluster 3.1.4, in which *LY6E* resides, it bears relation to cell signalling with by far the most significant and enriched term ‘chemotaxis’ (S3 Appendix). Chemotaxis is well known to play critical role in host response to infections, and is specifically involved in recruitment of leukocytes, and movement of lymphocytes around the body (55). The intersect of cluster with our model frequency analysis was also large, being 12 of its 40 genes *(ATF3, CCL2, CXCL10, HERC6, LAMP3, LGALS3BP, LY6E, OTOF, PARP12, SEPT4, SERPING1, and SIGLEC1*).

Cluster 3.1.5 contains genes involved in programmed cell death, containing several significant and enriched terms like ‘Apoptosis’, ‘Regulation of apoptotic process’ and ‘apoptotic process’ (S3 Appendix). A total of 3 of its 37 genes intersected our model frequency analysis (*CHMP5, FCGR1A, and FCGR1B*).

Illumina cluster 3.4 contained genes more related to general innate responses with enriched terms containing ‘Inflammatory response’ and ‘innate immune response’ with non-significant p-values (S3 Appendix). Amongst the genes are a number related to the Toll-like receptor family (also an enriched and significant term), which respond to microbial products and viruses, and are key-receptors of the innate immune system (56). Although not visible in the functional enrichment analysis, Illumina Cluster 3.4 also contained a number of Interleukin genes (*IL1B, IL1R1, IL4R, IL18R1, IRAK3*), known to be involved in inflammation and fundamental to innate immunity (57). Out of the 253 genes in cluster 3.4, 15, including CD177, intersected with previous frequency analysis (*BATF, CD177, DDAH2, GADD45A, GPR84, GRB10, GYG1, HK3, IRAK3, MAN2A2, MKNK1, NSUN7, SULT1B1, TSPO, and ZDHHC19*).

### Cross manufacturer gene list performance

We evaluated each of the BW & GA representative models from Affymetrix on the Illumina Data and Illumina Models on the Affymetrix data. Contrasting each model’s performance between these two discovery and non-discovery datasets we get the performance results depicted in Fig 5. This figure shows the difference between overall accuracy, and class-based accuracy, speciality and sensitivity when generalising our models to data pertaining from a different technology and set of studies.

**Fig 5.**
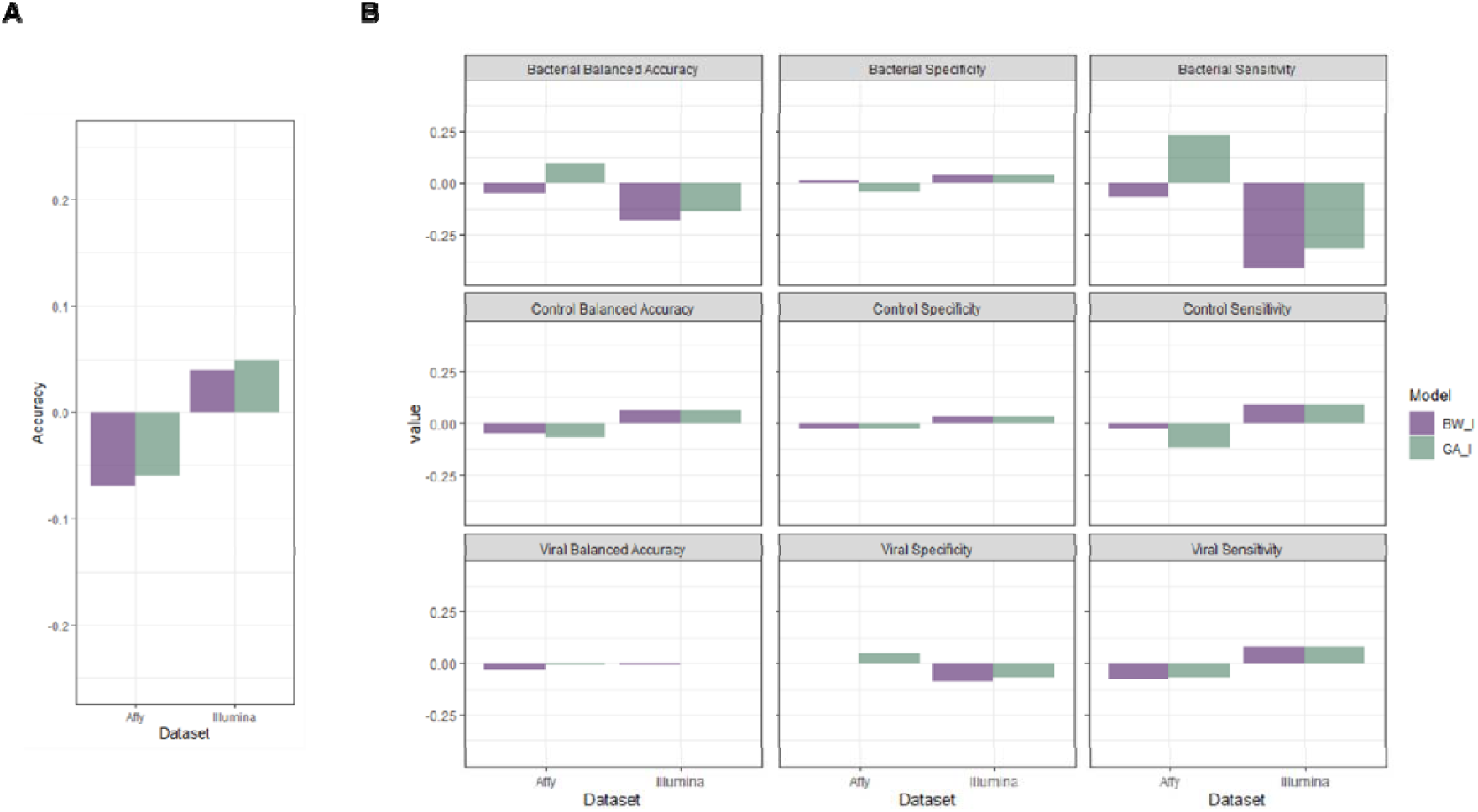
Cross manufacturer model change in performance. Difference in performance when taking Affymetrix derived models and testing on the Illumina data, and the Illumina derived models when testing on the Affymetrix data. (A) Difference in performance in terms of overall Accuracy. (B) Class based performance in terms of Balanced Accuracy, Sensitivity, and Specificity. For each performance measure, bars are grouped by model, and each bar refers to the difference between performance on the original dataset (which each model was discovered on) and the performance on the data it had not been exposed too. For Affymetrix models this would contrast the performance on the Affymetrix data,with the same model’s performance on the Illumina data.

In terms of overall accuracy (Fig 5 A) Affymetrix models, both GA ad BW, performed worse when applying to the Illumina data. However, the drop was less than 0.1 for both Affymetrix GA and BW. Whereas for Illumina, both GA and BW models slightly gained accuracy when applied to the Affymetrix data (0.04 and 0.05 respectively).

Looking specifically at bacterial performance (Fig 5 B), both Illumina models performed worse on the Affymetrix data in terms of bacterial balanced Accuracy (BW_I 0.71 and GA_I 0.73 2dp). Whereas the Affymetrix models performed well on the Illumina data (BW_I 0.89 and GA_I 0.89 2dp). In terms of bacterial specificity there was little change for all models, staying within +/-0.05 2dp of change in performance. However, in terms of bacterial sensitivity, the Illumina models performed particularly worse on the Affymetrix data (BW_I 0.44 and GA_I 0.47 2dp).

Across viral class specific metrics (Fig 5 B), no model had any large change in Balanced Accuracy (change < 0.05 2dp). The largest metric change was seen in sensitivity, with Affymetrix models slightly decreasing, but with an original score of 0.97 and 0.95 for BW_I and GA_I they are still performing well when ran on the Illumina data.

Overall, both Affymetrix and Illumina models performed well given that data was pertaining from different manufacturers and different groups of studies. Particularly stability around viral performance suggests a robustness to the gene lists for classifying viral samples correctly. However, given that bacterial performance change was very comparable to viral, it too suggests a strong ability to classify bacterial samples, even when moving out of the original dataset.

## Discussion

Due to the amount of relevant data, we focused our analysis on studies from two of the largest microarray platforms, Affymetrix and Illumina. Whilst these both determine the expression levels of genes and are common in large-scale population studies, differences in quantification and normalisation of gene expression values create technical difference (58). Studies within manufacturers were successfully batch corrected, indicated by non-significant changes in differentially expressed genes and removal of sample clustering by studies and platforms in PCA analysis. However, the combination of studies between manufacturers was unsuccessful, leading to two parallel analyses on the combined and batch corrected versions of (i) Affymetrix and (ii) Illumina datasets which minimized biological variation loss.

Simpler solutions are more specifically justifiable and allow for greater interpretation, which is the motivation for feature selection amongst models in biological data. We employed two feature selection algorithms using the Random Forest Classifier over our Data: Backwards Elimination and GALGO – both essentially cutting the noise and finding the most significant biological variation responsible for predicting disease state. It is unknown without a brute force search whether a *truly* optimal combination of genes has been found, however both BW and GA approaches converged around a small group of genes located in uncorrelated and functionally separable clusters. Models were found to be strongly enriched for the ISGs. In fact, *IFI27* and *LY6E* (both ISGs) were included in all Affymetrix and Illumina models. *IFI27* is involved in various signalling pathways affecting apoptosis (59-61). Whereas, *LY6E* belongs to a class of interferon-inducible factors that broadly enhance viral infectivity (62). *LY6E* has also been attributed a diverse set of effects, including attenuating T-cell receptor signalling (63) and suppressing responsiveness to *Lipopolysaccharide* which stimulate immune responses (64). Moreover, *IFI27* was shown by Tang et al. to be a *single–gene* biomarker that discriminates between influenza, and other viral and bacterial infections in patients with suspected respiratory infection (65). However, this single-gene biomarker approach lacks generalisability and robustness when predicting a more varied pathogen set. As we have observed, performance in our meta-analysis was greatly improved by including more genes in our models.

Our larger set of RF selected genes contained numerous examples confirmed by previous studies to be implicated in disease states. For instance, our results coincide with recent meta-analysis, by Andres-Terre et al., looking at transcriptional signatures of infections, specifically in distinguishing influenza from other viral and bacterial infections, which found 127 multi-gene signatures, 27 of which were also present in our representative models (*ATF3, BST2, CXCL10, EIF2AK2, HERC5, HERC6, IFI27, IFI44, IFI44L, IFI6, IFIT1, IFIT2, IFIT5, ISG15, JUP, LGALS3BP, LY6E, MRPL44, MTHFD2, MX1, OAS1, OAS2, OAS3, OASL, RSAD2, RTP4, SERPING1, SPATS2L*) serving to validate our successful data integration and biological findings (66). Notably amongst these coinciding genes are *IFI27* and *LY6E*, again confirming the validity of our converging feature selection.

By inferring the underlying interaction network, we discovered that convergence was not only happening to a set of genes, but also, and more prominently, convergence was focusing around particular groups of functionally similar genes. This gene-group convergence only emerged as part of an in-depth investigation into the driving forces of feature selection from a biological network perspective. When representative members of these uncorrelated gene clusters are taken together, they can form highly predictive gene lists. With the ability to define the host response to viral and bacterial infections, genes of our identified clusters are likely good at approximating key functions important in disease state prediction. Notably, the four functional groups of genes were indicated to be: Type I interferon-inducible genes (ISGs), Chemotaxis genes, Apoptotic Processes genes, and Inflammatory / Innate Response genes, which were prevalent in every model (both Affymetrix and Illumina). Within this cluster convergence we found a highly selected group of genes to be ISGs (the most frequent between both Affymetrix and Illumina models). This is no surprise, given Type I Interferons serve as a link between the innate and adaptive immune systems (67) and have a broad range of effects on both innate and adaptive immune cells during infection with viruses, bacteria, and parasites (47). Their varying sensitivity to particular forms of pathogens is likely why a number can be used in conjunction for classification with RFs. While ISGs exact function are not fully understood, it appears our RF models have identified their strong connection to disease state (68, 69). Whilst convergence was prominent around four functional groups of genes, we also note that both in Affymetrix and Illumina, a greater more variable set of functional gene groups were used in addition within our gene lists. Hence, there is a degree of variability in gene solutions, and it seems there is an interchangeable portion of our gene lists in which a number of genes from uncorrelated functional groups of genes can be used to achieve high performance in defining disease state.

Finally, we verified our gene lists for generalisability by retraining and evaluating on data from a different manufacturer to which they were discovered in (Affymetrix Gene lists to Illumina and Illumina Gene lists to Affymetrix). It is apparent that all gene lists tend to do better on Affymetrix data, regardless of which set they were discovered on, which suggests that the dataset, not the gene lists, is influencing performance. Hence, we have uncovered the differentiating biological signatures underlying able to define bacterial and viral infections.

## Conclusions

Our meta-analysis of Affymetrix and Illumina human blood infection data has revealed several panels of genes which are able to distinguish well between bacterial and viral infections. The difference in technology and gene coverage between Affymetrix and Illumina did not allow for a direct integration in our analysis. However, we were able to confirm that convergence was occurring independent of the technology, to both the same genes and the same functional groups of genes. This technology independent differentiable signal is learnable, and we demonstrated its presence by reconstructing the underlying regulatory gene network and overlaying models from the two datasets.

## Data Availability

All data is available at the following Gene expression Omnibus IDs: GSE49954, GSE50628, GSE54992, GSE25504, GSE66099, GSE69606, GSE6269, GSE18090, GSE28750, GSE34205, GSE52428,
GSE95104, GSE17156, GSE30550, GSE29385, GSE32707, GSE37250, GSE40396, GSE60244, GSE64456, GSE68310.

## Acknowledgments

We thank all the contributing studies for generating and making publicly available their respective datasets. We also gratefully acknowledge DSTL (www.gov.uk/dstl) for providing support.

This work was also supported by the Chem-Bio Diagnostics program contract HDTRA1-12-D-0003-0023 from the Department of Defense Chemical and Biological Defense program through the Defense Threat Reduction Agency (DTRA).

## Supporting information

**S1 Appendix. Pre-processing**.

**S1 Fig. Affymetrix Interaction Network**. Affymetrix recovered interaction network at first level of clustering. Selected model genes are highlighted.

**S1 Table. Model Gene Selection Frequency**. Affymetrix and Illumina model selected genes with relative frequency of selection (genes with greater than 5% aggregated inclusion across all search procedures).

**S2 Appendix. Biomarker Search**.

**S2 Fig. Illumina Interaction Network**. Illumina recovered interaction network at first level of clustering. Selected model genes are highlighted.

**S2 Table. Highly selected gene clusters from Affymetrix and Illumina interaction network**. Table containing the genes from the 4 highly model selected Illumina clusters, and 5 highly model selected gens from the Affymetrix clusters.

**S3 Appendix. Inferred Interaction Networks**.

**S3 Table. Out-sample results of gene lists**. The out-sample results from running Affymetrix derived gene lists on the Illumina data, and the Illumina derived gene lists on the Affymetrix data

